# Sorting out assortativity: when can we assess the contributions of different population groups to epidemic transmission?

**DOI:** 10.1101/2024.03.13.24304225

**Authors:** Cyril Geismar, Peter J White, Anne Cori, Thibaut Jombart

## Abstract

Characterising the transmission dynamics between various population groups is critical for implementing effective outbreak control measures whilst minimising financial costs and societal disruption.

Traditionally, mathematical models have primarily relied on assumptions of contact patterns to characterise transmission between groups. Thanks to technological and methodological advances, transmission chain data is increasingly available, providing information about individual-level transmission. However, it remains unclear how effectively and under what conditions such data can inform on transmission patterns between groups.

In this paper, we introduce a novel metric that leverages transmission chain data to estimate group transmission assortativity; this quantifies the extent to which individuals transmit within their own group compared to others. Through extensive simulations, we assessed the conditions under which our estimator performs effectively and established guidelines for minimal data requirements. Notably, we demonstrate that detecting and quantifying transmission assortativity is most reliable when groups have reached their epidemic peaks, consist of at least 30 cases each, and represent at least 10% of the total population each.

**Author Summary:** Efficient outbreak control relies on understanding how infection spreads between affected groups, such as healthcare workers and patients or specific age groups. Policies and interventions may differ substantially depending on how much transmission is within groups or between them. However, assessing transmission patterns between groups is challenging as these patterns are not only influenced by social contacts but also by variations in individual susceptibility and infectiousness, which changes over time. To address this challenge, we developed an estimator that utilises information on transmission chains (who infected whom), enabling the identification and quantification of transmission patterns between groups. Through extensive simulations, we assessed the conditions under which our estimator performs effectively and established guidelines for minimal data requirements. Our results suggest that inferring transmission patterns is most reliable when groups have reached their respective epidemic peaks, contain at least 30 cases each and constitute at least 10% or more of the total population, each.

## Introduction

In response to the COVID-19 pandemic, governments across the world implemented nationwide lockdowns, later transitioning to targeted pharmaceutical and non-pharmaceutical interventions based on factors such as location, age, and vaccination status. However, these measures could not always be optimised in real-time due to a lack of quantitative understanding regarding the varying roles different groups played in disease transmission. For example, the decision to close schools was initially based on the assumption that children were significant drivers of transmission [1,2]. Yet, subsequent research suggested that children may be less susceptible to infection and that schools may not have played a major role in the transmission of SARS-CoV-2 [3–6].

The ability to detect and quantify the contributions of different groups to transmission during an outbreak is essential for implementing effective control measures. Not only does it enhance our comprehension of transmission dynamics within a population, but it may also lead to better predictions of the epidemic’s future trajectory and enables the development of evidence-based public health strategies tailored to the outbreak’s characteristics.

Broadly, two approaches have been employed to assess the contributions of different groups to epidemic transmission. First, dedicated surveys have been conducted to measure the frequency of contact between different groups; combined with information about the relative infectiousness and susceptibility of each group (e.g. obtained from epidemiological or serological investigations), these data can be used by transmission models to estimate transmission assortativity [7,8]. Unfortunately, the underlying contact data can be biased, have limited sample size or representativeness, and may not be generalisable across different epidemic contexts [9–11].

Alternatively, transmission assortativity can be directly assessed from observed transmission patterns *e.g*. by measuring the proportion of cases in different groups [12,13] or by reconstructing the transmission chains [14,15]. These approaches have their own limitations. For instance, accurately reconstructing transmission chains is challenging [16] and even with perfectly known transmission chains, transmission assortativity estimation may be impeded by differences in group sizes and group-level saturation (*i.e*. the depletion of susceptibles).

This paper introduces a novel framework for evaluating transmission patterns among distinct groups during an outbreak, utilising known transmission chains to quantify group-specific assortativity. We evaluate the performance of our estimator through simulations across diverse outbreak scenarios and offer guidance on the minimum data collection requirements and the optimal estimation timeframe to inform policy.

## Methods

### A new estimator of transmission assortativity

Assortativity has been amply described for social mixing patterns, with homogeneous mixing referring to random contacts between individuals, and heterogeneous mixing denoting interactions characterised by distinct (non-random) patterns depending on group memberships [7]. Heterogeneous mixing can be either *assortative*, where individuals tend to interact more within their own group (*e.g*. social contacts by age [9,17,18]), or *disassortative*, where individuals interact preferentially with members of other groups (*e.g*. sexual contacts [19]). Here we use these definitions to characterise the patterns of transmission rather than contact.

To quantify transmission assortativity, we examine the person-to-person transmission patterns. We denote *β*_*b←a*_ the person-to-person transmission rate from an individual in group *a* to an individual in group *b*. We assume that *β*_*a←a*_ can be expressed as *β*_*a←a*_ = *γ*_*a*_ *β*_*b←a*_ (with *a* ≠ *b*), where *γ*_*a*_ is the assortativity coefficient for group *a. γ*_*a*_ is defined as the excess probability of a secondary infection taking place within group *a* compared to random expectation. *γ* values range from 0 (fully disassortative) to ∞ (fully assortative), with 1 indicating homogeneous patterns. For instance, *γ*_*a*_ = 2 indicates that an infected individual from group *a* is twice as likely to infect an individual from the same group compared to infecting an individual from another group. Conversely, a *γ*_*a*_ of 1/2 means that an infected individual from group *a* is twice as likely to infect an individual from another group compared to infecting an individual from the same group.

We consider *G* groups of relative sizes *f*_*1*_,…,*f*_*G*_ defined as:

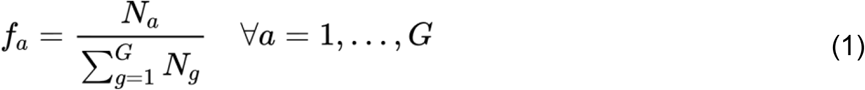

where *N*_*a*_ is the number of individuals in group *a*.

For an infectious individual in group *a*, the proportion *π*_*b←a*_ of secondary cases who are expected in group *b* is (details in supplementary materials S1.1):

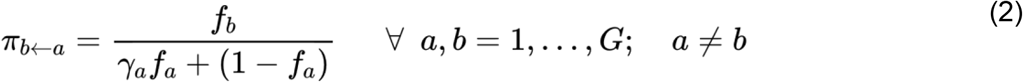

and

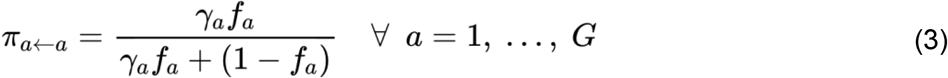

We can obtain *γ*_*a*_ by rewriting equation 3 as:

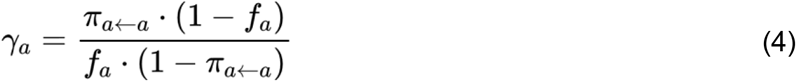

Here we assume that transmission chains are known. Among infections generated by infected individuals in group *a*, the proportion of secondary cases in group *a, π*_*a←a*_, can therefore be calculated as:

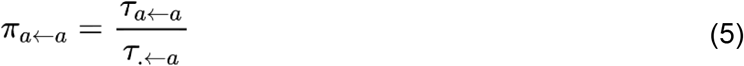

where *τ*_*a←a*_ is the number of observed within-group transmission pairs and *τ*_.*←a*_ is the total number of infections coming from group *a*. We can obtain a confidence interval (CI) on *π*_*a←a*_ for various significance (*α*) levels using the Clopper-Pearson binomial interval method [20] (S1.2). Feeding estimates of *π*_*a←a*_ from equation 5 into equation 4, provides estimates of *γ*_*a*_ with confidence intervals.

To simplify interpretation, we introduce a rescaled parameter *δ*, ranging between -1 (fully disassortative) and 1 (fully assortative), with 0 corresponding to a homogeneous transmission pattern (Figure S1), defined as:

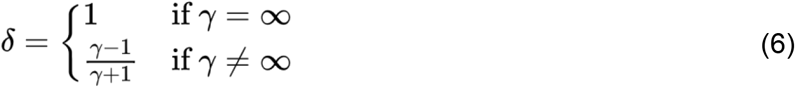

All our results are presented using *δ* rather than *γ*.

### Simulation study

We simulated numerous outbreaks under various contexts to assess the estimator’s performance. The simulation employed a discrete time branching process modelling individual infections spreading in successive generations. Simulations were specified with: i) group-level parameters including the size of each group, their assortativity coefficients (*δ*), initial introductions, basic reproduction numbers (*R*_0_) and ii) epidemic level parameters such as the number of groups, the pathogen generation time (*w*) and incubation period (*v*) distributions (both assumed the same across groups). The simulation outputs the transmission tree of the infected individuals including their group and that of their infector, their date of infection and date of symptom onset. We constructed 10,000 sets of input parameters, referred to as ‘scenarios’, by randomly sampling parameters from pre-defined distributions (S1.3, Figure S2). To account for stochasticity, we conducted 100 simulations for each unique scenario resulting in a total of 1,000,000 simulated outbreaks.

In our branching process model, the force of infection (FOI) generated by individual *j* from group *a* at time *t*, towards each individual in group *b* is defined as :

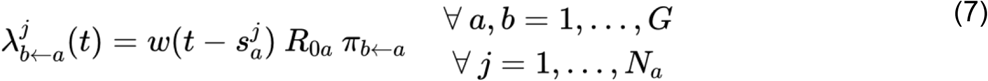

where:

- *s*^*j*^_*a*_ is the time of infection of individual *j* in group *a*
- *R*_0*a*_ is the basic reproduction number of individuals in group *a*

The total FOI that group *b* receives from all groups at time *t* is obtained as:

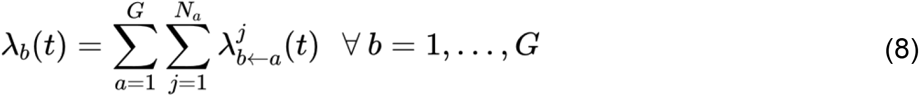

The probability of infection for each individual in group *b* at time *t* is then calculated as:

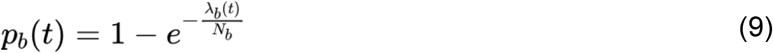

At time *t* + 1, the number of new cases in group *b, X*_*b*_(*t* + 1), is drawn from a binomial distribution:

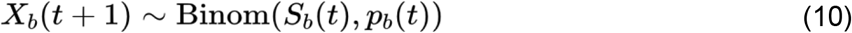

where *S*_*b*_(*t*) is the number of susceptible individuals in group *b* at time *t*.

New cases are allocated at random amongst susceptible individuals. The simulation progresses in discrete daily time steps for 365 days. Nearly all simulations (99.99%) finished with the last infection occurring before day 300. Note that we assume that individuals who have been infected become fully immune.

Assuming that *b*^*i*^ (i^th^ individual in group *b*) was infected at time *t*+1, their infector is drawn across all infected individuals in all groups from a multinomial distribution with probabilities:

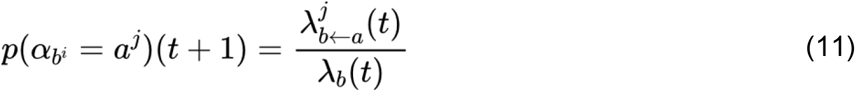

Where *a*^*j*^ is the *j*^*th*^ individual in group *a*.

To assess the performance of our estimator, we computed 4 different performance metrics for each scenario:

- *Bias*: defined as the average difference between the true *δ* value and its estimate 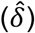 across 100 simulations. It is a measure of the estimator’s systematic error and inaccuracy and should be close to 0. Bias is positive when *δ* is underestimated, indicating underestimation of assortativity or overestimation of disassortativity. Conversely, negative bias occurs when *δ* is overestimated, indicating overestimation of assortativity or underestimation of disassortativity.
- *Coverage (at significance level α)*: defined as the proportion of simulations (out of 100) where the true *δ* value is within the estimated CI corresponding to *α*. We evaluate 4 significance levels 0.05, 0.1, 0.25 and 0.5. Assessing coverage helps determine the reliability of the confidence intervals generated by the estimator. Coverage should approximate 1-*α*, and the coverage error, which measures the deviation from this target, should be close to 0. A positive coverage error suggests underestimation of uncertainty, while a negative coverage error indicates overestimation.
- *Sensitivity* (true positive rate): defined as the proportion of simulations (out of 100) where the estimator correctly identifies a significant assortative or disassortative effect (i.e. the 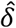 CI doesn’t contain 0). Sensitivity should be close to 1 (100%).
- *Specificity* (true negative rate): defined as the proportion of simulations (out of 100) where the estimator correctly identifies no significant assortative or disassortative effect (i.e. the 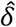 CI contains 0). Specificity should be close to 1 (100%).

We evaluated the estimator’s performance at various stages of the outbreak, defined in relation to the group’s epidemic peak, i.e. the day with the highest symptom onset incidence following the first case. Denoting *T* the date of the group’s peak incidence, we define the *analysis time window* as the time period from the first case of the group to day *T* x *ε*, where *ε* represents any non-negative real number and is referred to as the “peak coefficient”. A peak coefficient value of *ε=*1 implies analysis until the group’s peak, while values above or below 1 imply analysis using data up to before or after the peak respectively (S1.4, Figure S3). Additionally, we introduce the term ‘peak asynchronicity’, calculated as the standard deviation of peak dates *T* across groups, to measure heterogeneity in the groups’ peak dates.

To assess the impact of the scenario parameters on the performance metrics, separate regressions were conducted with each performance metric as a dependent variable and scenario parameters as independent variables (S1.5).

## Results

Figure 1 presents the estimator’s performance across all epidemic scenarios considered.

**Figure 1:**
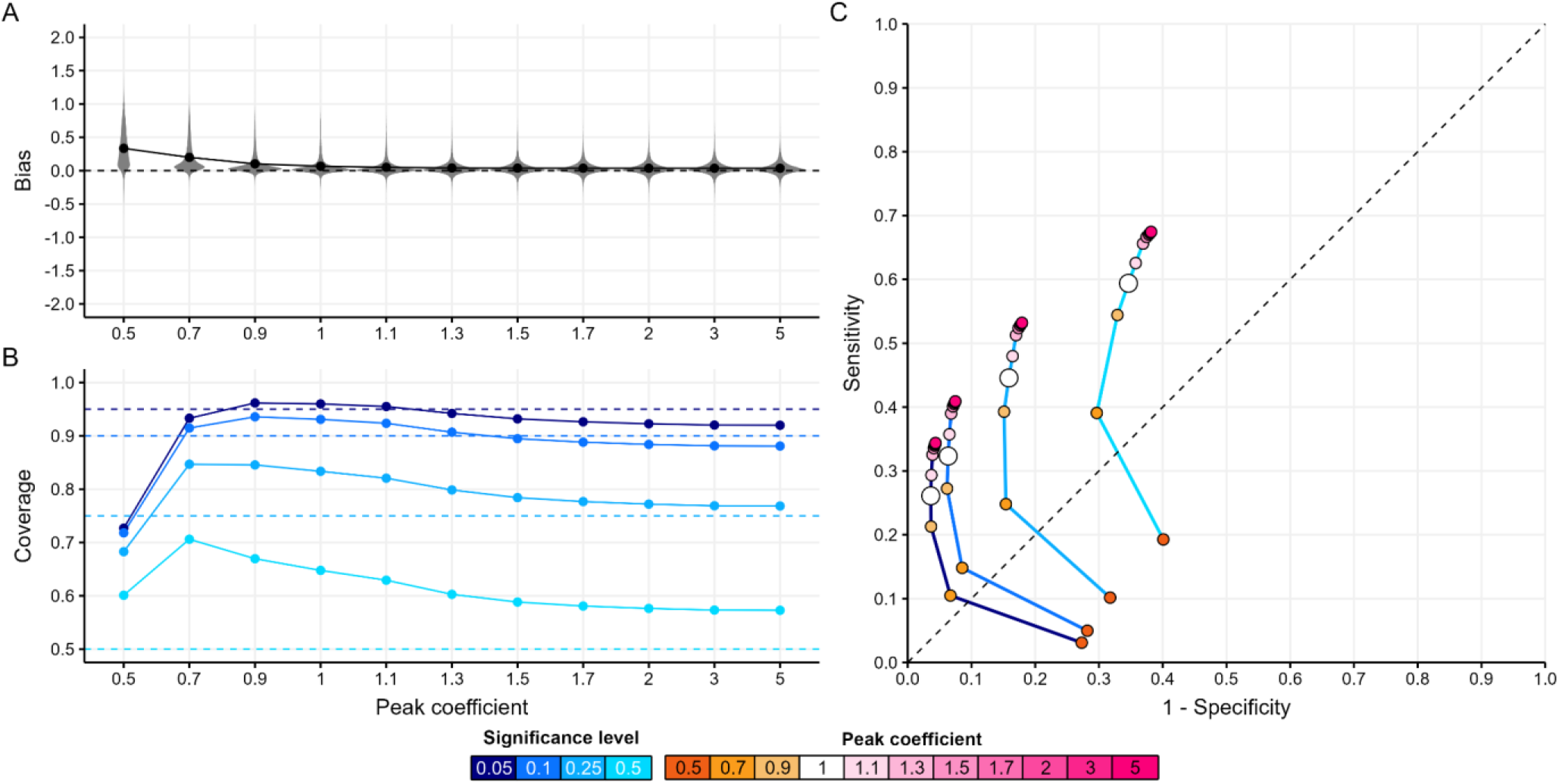
Estimator’s performance across all epidemic scenarios. A. Distribution of bias (the mean difference between the true assortativity *δ* value and its estimate) by peak coefficient. The peak coefficient (*ε*) is a non-negative real number used to define the *analysis time window* in relation to the group’s epidemic peak. It determines the analysis period from the first case to the day *Tε*, where *T* is the date of peak incidence for the group. A value of *ε*=1 indicates analysis up to the group’s peak date, while values above or below 1 extend the analysis to data after or before the group’s peak date, respectively. B. Mean coverage (proportion of simulations where the true *δ* value is within the estimated CI) by peak coefficient for each significance level (blue shades). C. The Receiver Operating Characteristic (ROC) (the trade-off between sensitivity and specificity) curves by peak coefficient (orange-pink points) for each significance level (blue shaded lines). In panel A, each point shows the mean metric value across all scenarios for a given peak coefficient. In panels B and C, each point shows the mean metric value across all scenarios for a given peak coefficient and significance level. Dashed lines refer to the metric’s target value for A and B and represent a random classifier’s ROC performance for C.

Bias decreased as the analysis time window expanded, achieving near-zero levels once the group had reached its epidemic peak (*ε=*1), with no substantial further improvements at later epidemic stages (*ε>*1, Figure 1A).

Coverage performance was contingent upon the significance (*α*) level and the stage of the group’s epidemic (*ε)* (Figure 1B). Halfway before the epidemic peak (peak coefficient *ε=0.5*), coverage at *α* levels up to 25% was too low, with average errors of 0.22, 0.18 and 0.07 for *α* levels of 5, 10, and 25%, respectively. In contrast, the 50% coverage was too high with an average error of -0.10. Around the epidemic peak (*ε* 0.7-1.3), coverage for *α* = 5-10% was good, whilst coverage for *α* = 25-50% was too high (average error -0.14). At later epidemic stages (*ε* 1.5-5), coverage was good across most significance levels, although the 50% coverage remained high across all epidemic stages.

Sensitivity and specificity were contingent upon the CI significance level *α* and the stage of the group’s epidemic (*ε*) (Figure 1C). Larger *α* values enhanced sensitivity at the expense of specificity, irrespective of the epidemic stage. And, regardless of *α*, analysing transmission chains later in the epidemic (i.e. increasing *ε*) also enhanced sensitivity, although this improvement was marginal past a peak coefficient of 1.5. However, the gain in sensitivity relative to the loss in specificity induced by delaying the analysis varied with *α*, with more pronounced tradeoffs for larger *α* values.

Figure 2 presents the relationship between various epidemic characteristics (columns) and the estimator’s performance metrics (rows), for a peak coefficient of 1 and a significance level of 0.05. Additional configurations are shown in supplementary materials (Figure S6).

**Figure 2:**
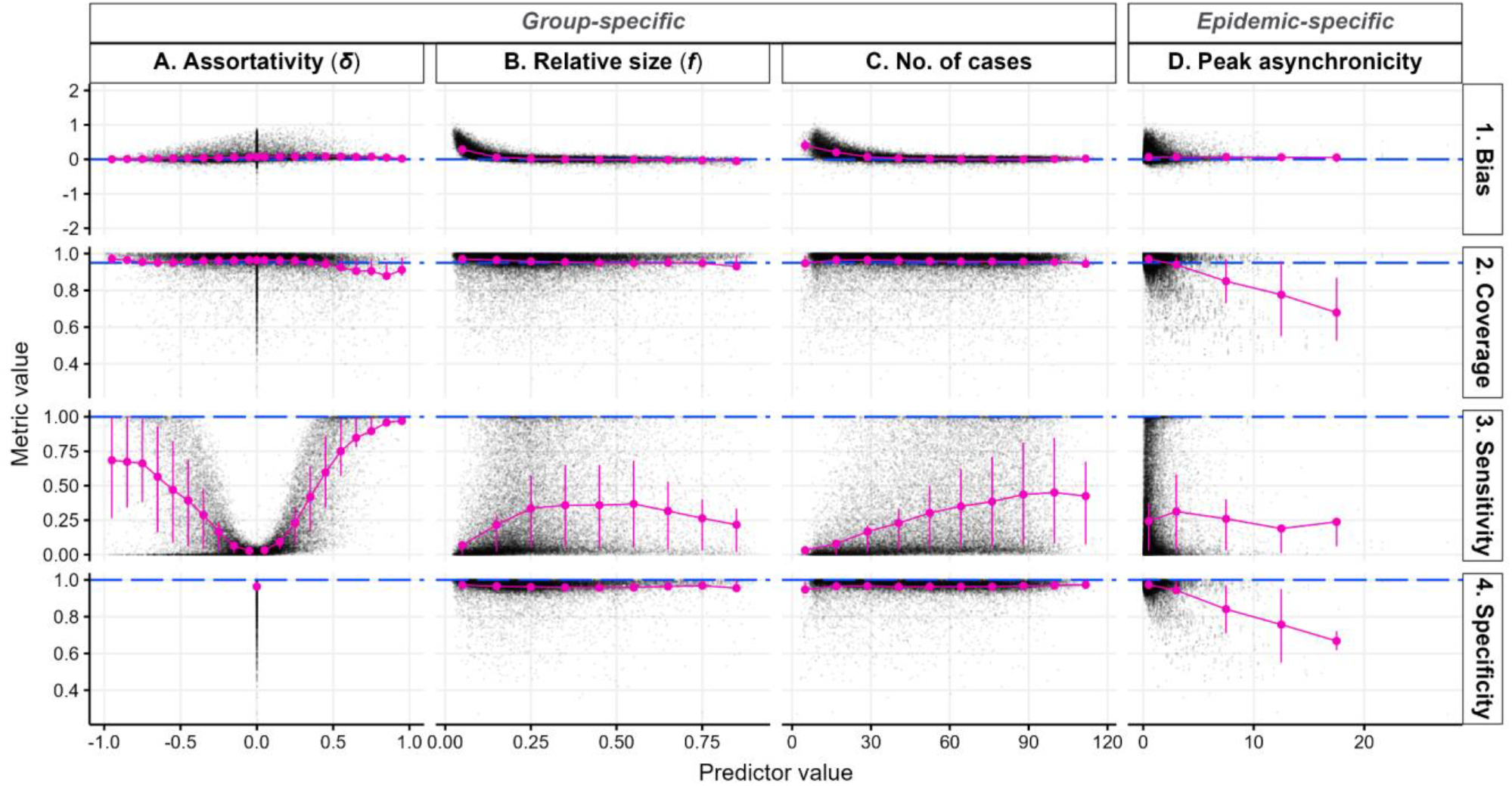
Estimator’s performance across scenario parameters and epidemic characteristics. Each row corresponds to one performance indicator and each column corresponds to one simulation parameter or epidemic characteristic. In each panel, the scatter plot depicts the univariate relationship between simulation parameter or epidemic characteristic (x-axis) and the performance metric (y-axis), where each black dot represents the average observation from 100 simulations for each group in every scenario. The pink points and error bars indicate the mean and interquartile range, calculated across different bin widths: 0.1 for *δ* (A.) and relative group size (B.), 12.5 for the number of cases in the group (C.) and 5 days for the standard deviation of peak date (D.). Dashed blue lines indicate target metric values. Transmission chains were analysed up to the group’s epidemic peak (*ε*=1), with a significance level of 0.05.

Our estimator maintained consistent unbiased performance across the entire assortativity range (*δ* from -1 to 1) (Figure 2 column A row 1). Coverage consistently met the 95% target for *δ <* 0.5, with a slight decrease in coverage performance for *δ* > 0.5, although coverage remained close to the target, averaging at 0.91 (sd = 0.10) (Figure 2A2). This decrease in coverage in highly assortative scenarios could be due to a saturation effect: high assortativity will accelerate the depletion of susceptibles in the group, eventually resulting in lower observed assortativity compared to the true value (Figure S4). Although the assortativity coefficient *δ* only had a small effect on bias or coverage, it had a substantial impact on sensitivity, which was higher for larger absolute values of *δ*. However, sensitivity rose more gradually as |*δ*| increased on the disassortative scale compared to the assortative scale (Figure 2A3, Table S1.1), reaching an average of 82% for *δ* ≥ 0.5 compared to 55% for *δ ≤* -0.5, suggesting a better ability to detect assortative than disassortative transmission. Indeed, assortative transmission implies that transmissions propagate within the same group across multiple generations, consequently increasing the sample size (*τ.←a* in equation 5) compared to disassortative transmission, and thus narrowing the CI, thereby enhancing sensitivity. Our linear regression suggested that the assortativity coefficient explained nearly 60% of the variance observed in sensitivity (Table S1.1).

Increasing the number of cases substantially reduced bias (Figure 2C1, Table S2), and increased sensitivity (Figure 2C3, Table S1.2) but had little effect on specificity or coverage (Figure 2C4 and 2C2). Bias was negligible (mean: 0.04, sd: 0.07) once the group reached 30 to 40 cases. Sensitivity was positively correlated with the number of cases: controlling for *δ*, the odds of detecting an assortative or disassortative pattern increased by 4% with each additional case (Table S1.2).

The relative size of the group had a substantial effect on bias (Figure 2B1, Table S2) and sensitivity (Figure 2 B3, Table S1.2) but no effect on specificity (Figure 2B4) nor coverage (Figure 2B2). When groups comprised 10% or more of the total population size, bias was close to 0 (Figure 2B1), and the odds of detecting an assortative pattern increased fourfold, compared to smaller groups (odds ratios (OR) = 4.15, 95% CI = 4.07 – 4.24) (Figure 2B3, Table S.1.2). Relative size and the number of cases jointly accounted for 72% of the variation in bias (Table S2), and contributed to a 42% increase in the pseudo R-squared for the linear regression on sensitivity (from 0.566 in Table S1.1 to 0.805 in Table S1.2).

Diverse transmission dynamics emerge from numerous groups, varying group sizes, reproduction numbers, and/or assortativity coefficients (Figure S5). This diversity results in varying saturation levels between groups over time, affecting transmission patterns within and between groups. Peak asynchronicity, a measure of heterogeneity in epidemic peak timing across groups was negatively associated with coverage (OR = 0.78, 95% CI = 0.78-0.78) and specificity (OR = 0.76, 95% CI = 0.76-0.76), explaining 18% and 24% of the variance, respectively (Table S3 and S4, Figure 2D2 and 2D4). These results suggest a decrease in our estimator’s performance with increasing heterogeneity between groups. However, our estimates remained unbiased (Figure 2D1) and with consistent sensitivity (Figure 2D3) irrespective of that heterogeneity.

In summary, analysing transmission chains at least up to the group’s epidemic peak generally improved all performance metrics. Near the group’s epidemic peak, coverage with significance levels of 5 or 10% yielded good performance, while levels of 25 and 50% were a bit too high, improving after the peak. Specificity was higher at lower significance levels, while sensitivity was higher at larger significance levels. Increased cases and relative group size contributed to improved estimator accuracy, reduced bias, and heightened sensitivity, with no significant impact on coverage nor specificity. Complex epidemic settings, measured through peak asynchronicity, did not significantly affect sensitivity or bias but were associated with a reduction in coverage and specificity.

## Discussion

We developed a method to detect and quantify the transmission assortativity of different groups based on transmission chains. We performed an extensive simulation study covering a range of epidemic scenarios to assess the performance of our approach.

Our results indicate that the estimator’s performance is influenced by assortativity patterns, relative group sizes, number of cases, and peak dates asynchronicity.

Generally, analysing transmission chains too early in the outbreak, before the group’s epidemic peak, results in poor performance across all metrics considered. On the other hand, delaying assortativity coefficient estimation poses challenges for timely policy implementation. Choosing when exactly in the epidemic to analyse transmission chains, and what significance level to use for estimating the assortativity coefficients, will also depend on the objective. For instance, minimising bias and maximising sensitivity is best achieved later in the epidemic, past the group’s peak, and using larger significance levels. Conversely, improving coverage and maximising specificity is easiest before the group’s epidemic peak and using lower significance levels. Nevertheless, estimating assortativity at a target time before or at the peak requires accurate prediction of the group’s peak date which can be very challenging.

As a rule of thumb, we suggest analysing all available transmission chain data up to the group’s epidemic peak with a significance level of 0.05. Under this setting, our estimator provides a generally accurate measure of assortativity with reliable coverage and specificity albeit lower sensitivity.

Detecting non-homogeneous transmission patterns (sensitivity) in the presence of relatively small groups (*i.e*. a group constituting less than 10% of the total population), with groups having fewer than 30 cases is challenging, particularly when assortative or disassortative patterns are mild (−0.5 ≤ *δ* ≤ 0.5). Importantly, it is considerably easier to detect assortativity than disassortativity, given that assortativity yields more transmission events within the group considered (where most new infections appear) compared to disassortativity (where new infections tend to appear in other groups, by definition). Hence, all other things being equal, larger sample sizes are more easily achieved in assortative groups.

Our approach complements traditional survey-based methods when transmission chains are available. Worby *et al*.’s relative risk estimation [12], measuring each group’s proportional change in infection incidence before and after the peak, and Abbas *et al*.’s assessment method [15], comparing actual and expected proportions of infections across groups, do not consider the influence of group size. By integrating group size into our approach, we account for variations in the pool of susceptible individuals within each group, offering a more comprehensive understanding of transmission dynamics. Consequently, our approach should provide novel insights into the impact of group dynamics when estimating transmission patterns.

The main limitation of our approach pertains to the assumption that transmission chains are perfectly known. Although transmission trees can be reconstructed from data, such reconstruction effort comes with inherent uncertainty, which we have not considered here. Conventional epidemiological investigations may provide reliable transmission chains but require intensive labour for contact tracing, data collection and analysis, and may be prone to error [21]. Statistical approaches have been developed to reconstruct who infected whom using data on contacts, symptoms onset dates, and pathogen genome sequences [22], but in some contexts even these prove insufficient to precisely reconstruct transmission trees [14,23]. Our study underscores the challenges of inferring group contributions in some scenarios, even in the hypothetical instance where transmission trees are perfectly known. Nevertheless, our approach is adaptable and can be extended to reconstructed transmission chains, for example, by estimating the assortativity coefficient across all posterior transmission trees in the setting of Abbas *et al*. [15]. Future research should delve into understanding how uncertainty surrounding these transmission trees further impacts our ability to infer transmission patterns.

Another limitation of our approach includes that our estimator requires information on group sizes which may be difficult to obtain in real-life settings, however various methods exist for population size estimation [24]. Our simulations also assumed that individuals who have been infected become permanently immune, an assumption which is typically valid over short time frames but may be unrealistic over longer time horizons.

Despite these limitations, this study provides a valuable first step towards evaluating the contributions of different groups to the transmission of infectious diseases and informing targeted control policy.

## Data Availability

The analysis code is freely available on a GitHub repository: https://github.com/CyGei/o2groups-analysis. An R package has been developed for simulating outbreak scenarios and is also available on GitHub at: https://github.com/CyGei/o2groups.

Package and analysis code have been archived on Zenodo (analysis: https://zenodo.org/doi/10.5281/zenodo.10616176, package: https://zenodo.org/doi/10.5281/zenodo.10616155)

## Acknowledgements

CG is supported by a PhD studentship at Imperial College London funded by the National Institute for Health Research (NIHR) Health Protection Research Unit (HPRU) in Modelling and Health Economics, which is a partnership between the UK Health Security Agency (UKHSA), Imperial College London, and the London School of Hygiene & Tropical Medicine (grant code NIHR200908). AC, PJW are supported by the HPRU in Modelling and Health Economics. This work was supported by the UK Medical Research Council (MRC) Centre for Global Infectious Disease Analysis (grant number MR/X020258/1); this award comes under the Global Health EDCTP3 Joint Undertaking.

## Author Contributions

Conceptualization: CG, AC, TJ, PJW.

Methodology: CG, AC, TJ, PJW.

Software: CG.

Validation: AC, TJ, PJW, CG.

Formal analysis: CG, AC, TJ, PJW.

Data Curation: CG.

Original Draft: CG.

Writing: CG, TJ, PJW, AC.

Visualisation: CG.

Supervision: AC, TJ, PJW.

## Notes

### Competing Interest Statement

The authors have declared no competing interest.

