## Supplementary material for "Sorting out assortativity: when can we assess the contributions of different population groups to epidemic transmission?"

The following presents the supplementary materials for the paper entitled: "*Sorting out assortativity: when can we assess the contributions of different population groups to epidemic transmission?*" The first part provides supplementary materials related to the methodology. The second part covers additional results.


1. Methodology 2
   1.1. Deriving $\pi_{b\leftarrow a}$ from the person-to-person transmission rate and relative group size. 2
   1.2. Calculating the confidence interval for $\pi_{a\leftarrow a}$ from observed transmission chains. 3
   1.3. Epidemic scenario parameterisation 5
   1.4. Peak coefficient ε 7
   1.5. Model development and evaluation framework 8
2. Additional Results 10

3. References 17

### 1. Methodology

#### 1.1. Deriving $\pi_{b \leftarrow a}$ from the person-to-person transmission rate and relative group size.

$\beta_{b \leftarrow a}$ denotes the person-to-person transmission rate from an individual in group *a to* an individual in group *b.* It follows that, in a fully susceptible population, the expected number of secondary cases in group *b* generated by one individual in group *a* is proportional to $\beta_{b \leftarrow a} N_b \propto \beta_{b \leftarrow a} f_b$, where $N_b$ is the number of individuals in group *b* and $f_b$ is the relative size of group *b* (equation 1 in the main text). We assume that $\beta_{b \leftarrow a}$ is the same for all $b \neq a$, i.e. $\beta_{b \leftarrow a} = \psi$ if a ≠ b, and $\beta_{a \leftarrow a} = \gamma_a \psi$. $\gamma_a$ is the assortativity coefficient for group *a* and is defined as the excess probability of a secondary infection taking place within group *a* compared to random expectation. Based on these assumptions, we derive the proportion of secondary cases who are in group *b* among the cases generated by an infectious individual in group *a,* $\pi_{b \leftarrow a}$, as:

$$\pi_{b \leftarrow a} = \frac{\beta_{b \leftarrow a} f_b}{\sum_{g=1}^{G} \beta_{g \leftarrow a} f_g} = \frac{\beta_{b \leftarrow a} f_b}{\sum_{\substack{g=1 \\ g \neq a}}^{G} \beta_{g \leftarrow a} f_g + \beta_{a \leftarrow a} f_a} = \frac{\beta_{b \leftarrow a} f_b}{\psi(1 - f_a) + \gamma_a \psi f_a}$$

<u>Hence:</u>

$$\pi_{a \leftarrow a} = \frac{\beta_{a \leftarrow a} f_a}{\psi(1 - f_a) + \gamma_a \psi f_a} = \frac{\gamma_a \psi f_a}{\psi(1 - f_a) + \gamma_a \psi f_a} = \frac{\gamma_a f_a}{(1 - f_a) + \gamma_a f_a}$$

<u>And if *a* ≠ *b*:</u>

$$\pi_{b \leftarrow a} = \frac{\beta_{b \leftarrow a} f_b}{\psi(1 - f_a) + \gamma_a \psi f_a} = \frac{\psi f_b}{\psi(1 - f_a) + \gamma_a \psi f_a} = \frac{f_b}{(1 - f_a) + \gamma_a f_a}$$

#### 1.2. Calculating the confidence interval for $\pi_{a\leftarrow a}$ from observed transmission chains.

Equation 5 in the main text is obtained by calculating $\pi_{a\leftarrow a}$ as the ratio between $\tau_{a\leftarrow a}$ and $\tau_{.\leftarrow a}$. Confidence intervals around that mean estimate can be obtained using the Clopper-Pearson binomial interval method [1], as: $\pi_{a\leftarrow a_{\text{lb}}} = Beta\left(p = \frac{\alpha}{2}; s1 = \tau_{a\leftarrow a}, s2 = \tau_{.\leftarrow a} - \tau_{a\leftarrow a} + 1\right)$

$$\pi_{a\leftarrow a_{\text{ub}}} = Beta\left(p = 1 - \frac{\alpha}{2}; s1 = \tau_{a\leftarrow a} + 1, s2 = \tau_{.\leftarrow a} - \tau_{a\leftarrow a}\right)$$

where*:*

- $\pi_{a\leftarrow a\ lb}$ is the lower bound of the CI for $\pi_{a\leftarrow a}$.
- $\pi_{a\leftarrow a\ ub}$ is the upper bound of the CI for $\pi_{a\leftarrow a}$.
- *Beta* is the quantile function of the beta distribution.
- *p* is the probability value.
- $\alpha$ is the significance level.
- *s1* and *s2* are the shape parameters of the *Beta* distribution.
- $\tau_{a\leftarrow a}$ is the number of within-group transmissions in group *a*.
- $\tau_{.\leftarrow a}$ is the total number of transmissions emitted from group *a*.

Note that the relationship between $\gamma_a$ and $\pi_{a\leftarrow a}$ in equation 6 of the manuscript, is monotonic and hence preserves confidence intervals.

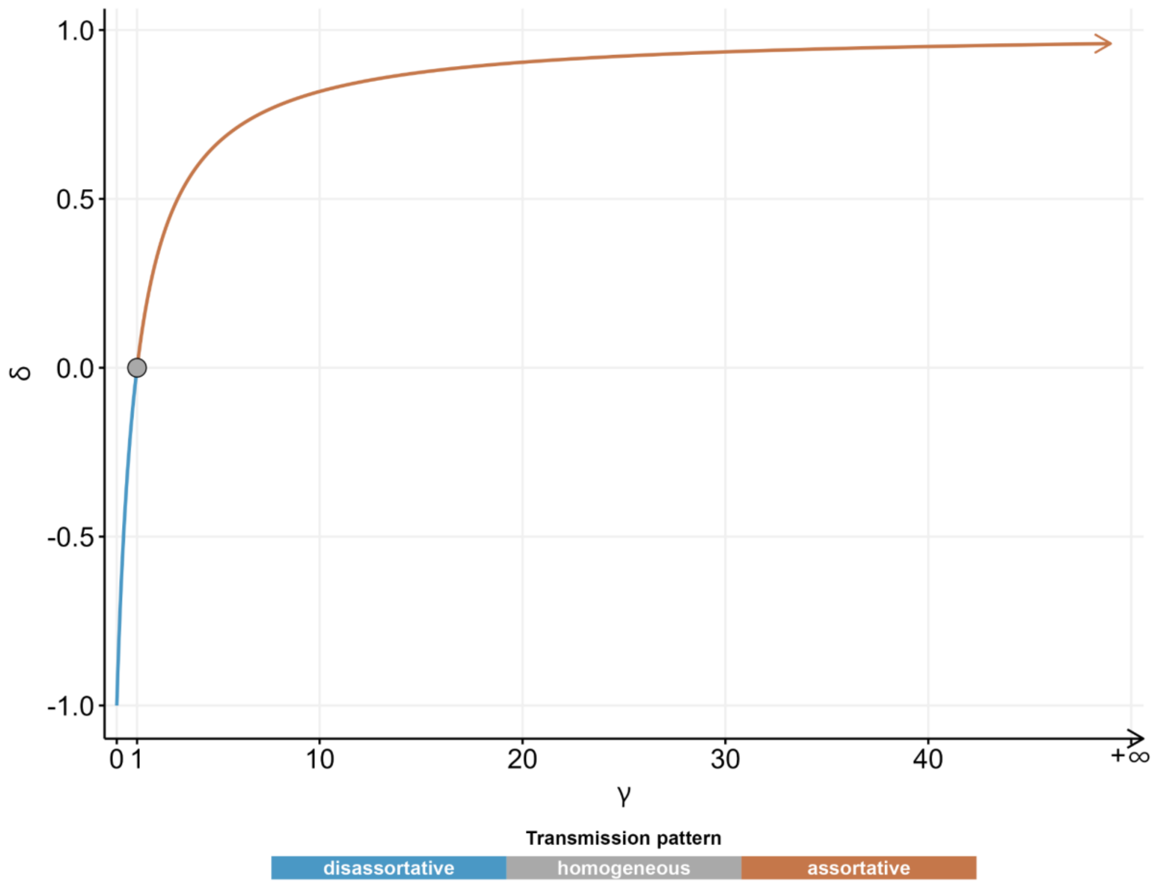


**Figure S1: Relationship between *δ* and *γ* as defined by equation 6 of the manuscript.**

Blue, grey and brown refer to disassortative, homogeneous (random) and assortative transmission patterns, respectively.

##### 1.3. Epidemic scenario parameterisation

- **Number of groups**: drawn from a truncated normal distribution with a mean of 2.5 and a standard deviation of 3. The truncation bounds are set to 2 and 8 (included). The resulting value is rounded to the nearest integer.
- **Group size**: values for each group are drawn independently from a uniform distribution between 20 and 200. Each group's size is rounded to the nearest integer.
- **Assortativity coefficient**: each group's $\delta$ value is drawn as follows:
  - $\delta = 0$ (homogeneous transmission) with probability 50%
  - Conditional on not being zero, $\delta$ is drawn from a truncated normal distribution with a mean of 0 and a standard deviation of 0.35, bounded between -1 and 1, thereby ensuring equal probability for each group to be assigned either a dis/assortative or homogeneous transmission coefficient.
- **Number of introductions**: an introduction denotes the initial occurrence of an individual contracting an infection. Initially, one group is randomly chosen to experience a single introduction. Following this, each group receives extra introductions, randomly drawn from a uniform distribution ranging between 0 and 10% of the group's population size.
- **$R_0$**: values for each group are drawn from a truncated normal distribution with a lower bound of 1, mean of 2, and a standard deviation of 2.
- **Natural histories**: The mean ($\mu$) and standard deviation ($\sigma$) for the natural histories are drawn from truncated normal distributions with lower bounds of 1, means of 4, and standard deviations of 3 (all measured in days). The generation time (GT) and incubation period (INCUB) distributions are then modelled using discretised gamma distributions.

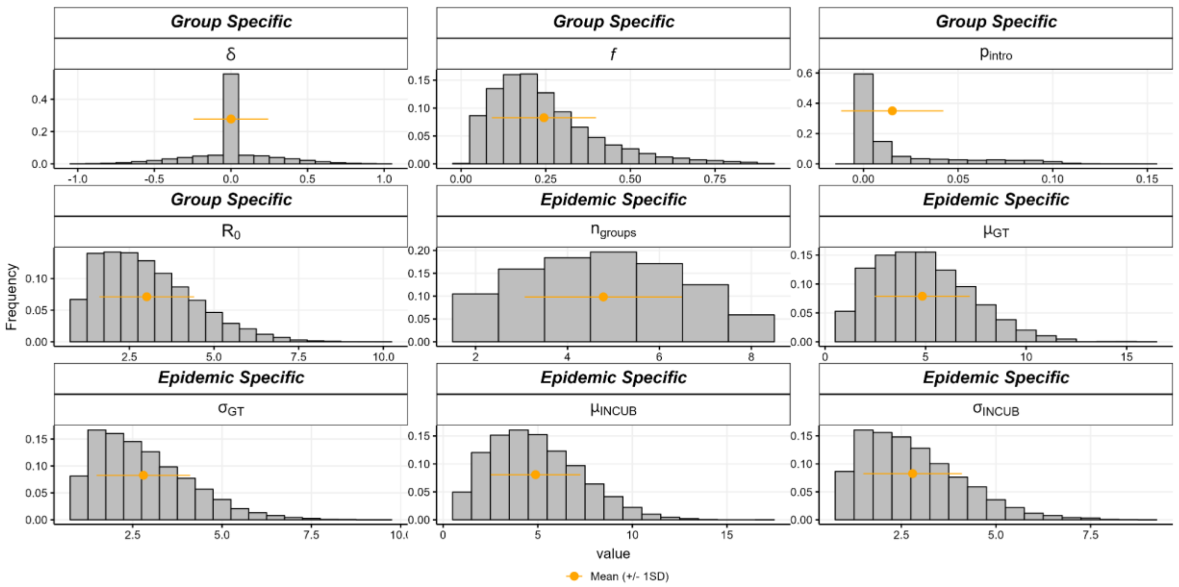


**Figure S2: Frequency histograms of scenario parameters**

Panel headers labelled 'Group Specific' indicate parameters drawn for each individual group, while 'Epidemic Specific' headers denote parameters drawn at the scenario level. Labels are defined as follows:

- δ : the assortativity coefficient.
- *f*: proportion of the population belonging to the group.
- $R_0$: basic reproduction number.
- $p_{intro}$: the proportion of initial introductions relative to the group size.
- $n_{groups}$: the number of groups.
- $\mu_{GT}$: the mean of the generation time distribution (in days).
- $\mu_{INCUB}$: the mean of the incubation period distribution (in days).
- $\sigma_{GT}$: the standard deviation of the generation time distribution (in days).
- $\sigma_{INCUB}$:  the standard deviation of the generation time distribution (in days).

In each panel, yellow points and error bars represent mean +/- one standard deviation.

##### 1.4. Peak coefficient ε

The peak coefficient ($\varepsilon$), is a non-negative real number used to define the *analysis time window* in relation to the group's epidemic peak. It determines the analysis period from the first case to the day $T\varepsilon$, where $T$ is the date of peak incidence for the group. A value of $\varepsilon$=1 indicates analysis up to the group's peak date, while values above or below 1 extend the analysis to data after or before the group's peak date, respectively. For example, if the peak date for the group of interest is on day 10, analysing transmission chains at a peak coefficient of ε=1.3 signifies investigating all transmissions up to day 13 (10 * 1.3 = 13).

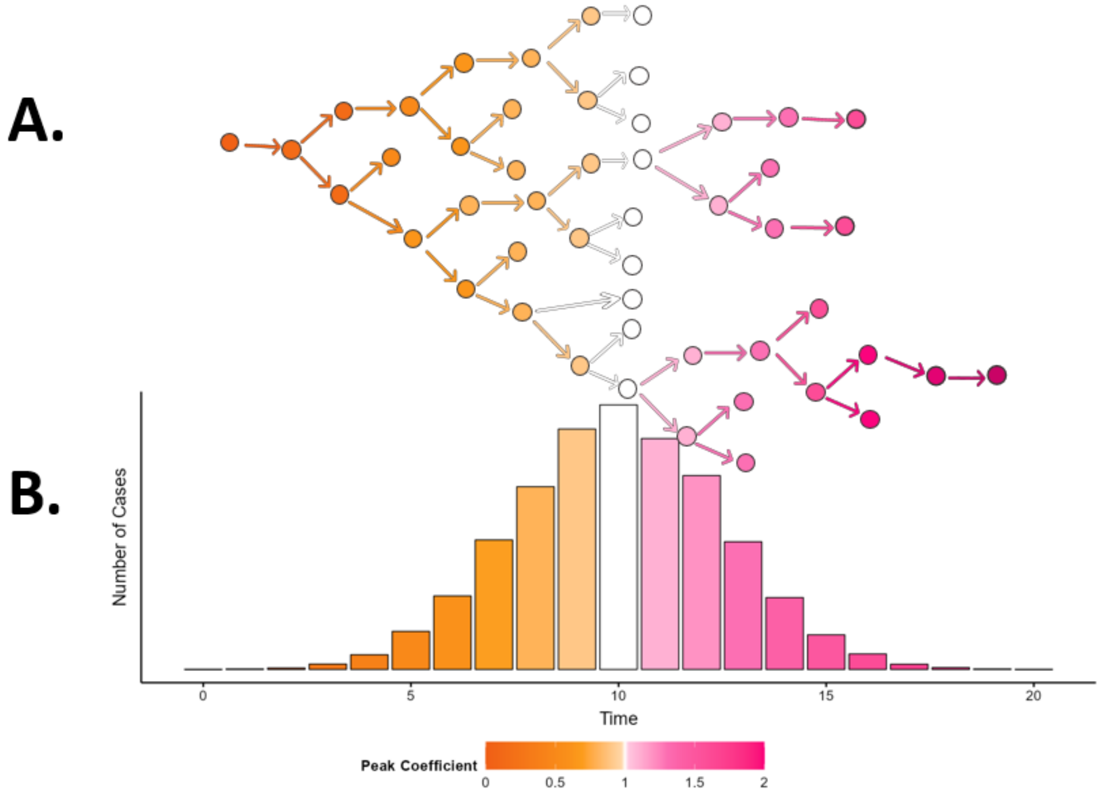


**Figure S3: Relationship between the epidemic transmission tree (A.), the group's epidemic curve (B.) and the group's peak coefficient (colour).**

In panel A, the schematic transmission tree depicts all cases (irrespective of group) as nodes, with edges representing the direction of transmission. The tree is ordered by the dates of symptom onset (x-axis in panel B). The epidemic curve in panel B shows the number of cases in a given group over time. In both panels, colours denote peak coefficient values using the same colour scheme as in Figure 1.

#### 1.5. Model development and evaluation framework

We employed 10-fold cross-validation to evaluate different regression models for predictor selection. The selection process involved identifying the best-performing models based on the highest R-squared values (for linear regressions) and pseudo (McFadden [2]) R-squared values (for logistic regressions) for each outcome. Subsequently, the models identified through cross-validation were fitted to the entire dataset, and the resulting regression coefficients along with model performance metrics were reported in section 2. The formulations for the final selected models are presented below. Note that, to assess the variance in sensitivity solely explained by the assortativity coefficient, we performed the logistic regression expressed in 1.1.

$$logit(Sensitivity) = \beta_0 + \beta_1 \mathbf{1}_{\{\delta_{<0}\}} + \beta_2|\delta| + \beta_3 \mathbf{1}_{\{\delta_{<0}\}} \cdot |\delta| + \epsilon \quad (1.1)$$

$$logit(Sensitivity) = \beta_0 + \beta_1 \mathbf{1}_{\{\delta_{<0}\}} + \beta_2|\delta| + \beta_3 n_{cases} + \beta_4 \mathbf{1}_{\{\delta_{<0}\}} \cdot |\delta| + e \quad (1.2)$$

$$Bias = \beta_0 + \beta_1 \log(f) + \beta_2 \log(n_{cases}) + \beta_3 \mathbf{1}_{\{f \geq 0.10\}} + \beta_4 \mathbf{1}_{\{n_{cases} \geq 30\}} + \beta_5 \log(f) \cdot \mathbf{1}_{\{f \geq 0.10\}} + \beta_6 \log(n_{cases}) \cdot \mathbf{1}_{\{n_{cases} \geq 30\}} + e \quad (2)$$

$$logit(Coverage) = \beta_0 + \beta_1 \sigma_{peak} + e \quad (3)$$

$$logit(Specificity) = \beta_0 + \beta_1 \sigma_{peak} + e \quad (4)$$

- Here, $\beta$ refers to the regression coefficient for the associated predictor.
- $\delta$ refers to the assortativity coefficient of the group.
- $|\delta|$ refers to the absolute value of the group's assortativity coefficient.
- $\mathbf{1}\{\delta_{<0}\}$ refers to a categorical variable indicating whether the group is disassortative or assortative.
- $n_{cases}$ refers to the number of cases in a given group.
- $\mathbf{1}\{n_{cases \geq 30}\}$ refers to a categorical variable indicating whether the group had at least 30 cases.
- $f$ refers to the relative size of the group *i.e.* the proportion of the total population belonging to the group.

- $\mathbf{1}\{f_{\geq 0.10}\}$ refers to a categorical variable indicating whether the group represented 10% or more of the total population.
- $\sigma_{\text{peak}}$ refers to the standard deviation in the peak dates across groups, also referred to 'peak asynchronicity' in the manuscript.
- $|\delta|$ refers to the absolute value of the assortativity coefficient for the group.
- log represents the natural logarithms base 10 transformation of the given variable.
- logit represents the log-odds transformation used in logistic regression.
- $e$ represents the residual errors in the multivariate linear regression model.

The logistic regressions (equations 1.1, 1.2, 3, 4) were fitted by Maximum-Likelihood (ML) method for parameter estimation, and their results presented in the tables in section 2. Odds ratios (2$^{nd}$ column), along with 95% confidence intervals (3$^{rd}$ column), and p-values (4$^{th}$ column) were reported. The last two rows report the number of observations and the pseudo (McFadden [2]) R-squared.

The linear regression (equation 2) was performed on values of bias and fitted by Maximum-Likelihood (ML). ML estimates are indicated in the second column, with associated 95% confidence intervals (3rd column) and p-values (4th column). The last two rows report the number of observations and the R-squared

### 2. Additional Results

**Table S1: (1.1) Multivariable model of sensitivity (equation 1.1 of section 1.5).**

| ***Predictors*** | ***Odds ratios*** | ***Confidence interval*** | ***p-value*** |
|---|---|---|---|
| (Intercept) | 0.03 | 0.03 – 0.03 | **<0.001** |
| $\mathbb{1}\{\delta_{<0}\}$ | 1.30 | 1.27 – 1.32 | **<0.001** |
| $\lvert\delta\rvert$ | 4990.48 | 4822.32 – 5164.87 | **<0.001** |
| $\mathbb{1}\{\delta_{<0}\} \times \lvert\delta\rvert$ | 0.06 | 0.05 – 0.06 | **<0.001** |
| Observations | 20478 | | |
| McFadden $R^2$ / McFadden $R^2$ adjusted | 0.566 / 0.566 | | |

**Table S1.2: Multivariable model of sensitivity (equation 1.2 of section 1.5)**

| ***Predictors*** | ***Odds ratios*** | ***Confidence interval*** | ***p-value*** |
|---|---|---|---|
| (Intercept) | 0.00 | 0.00 – 0.00 | **<0.001** |
| $\mathbb{1}\{\delta_{<0}\}$ | 1.46 | 1.44 – 1.49 | **<0.001** |
| $\lvert\delta\rvert$ | 31552.94 | 30305.12 – 32855.00 | **<0.001** |
| $n_{cases}$ | 1.04 | 1.04 – 1.04 | **<0.001** |
| $\mathbb{1}\{f_{\geq 0.10}\}$ | 4.15 | 4.07 – 4.24 | **<0.001** |
| $\mathbb{1}\{\delta_{<0}\} \times \lvert\delta\rvert$ | 0.03 | 0.03 – 0.03 | **<0.001** |
| Observations | 20478 | | |

| | |
|---|---|
| McFadden $R^2$ / McFadden $R^2$ adjusted | 0.805 / 0.805 |

**Table S2: Multivariable model of bias (equation 2 of section 1.5)**

| ***Predictors*** | ***Estimates*** | ***Confidence interval*** | ***p-value*** |
|---|---|---|---|
| (Intercept) | -0.10 | -0.12 – -0.07 | **<0.001** |
| **log($f$)** | -0.29 | -0.30 – -0.29 | **<0.001** |
| $\mathbf{1}\{f_{\geq 0.10}\}$ | 0.55 | 0.53 – 0.56 | **<0.001** |
| **log($n_{cases}$)** | -0.15 | -0.15 – -0.15 | **<0.001** |
| $\mathbf{1}\{n_{cases \geq 30}\}$ | -0.48 | -0.50 – -0.47 | **<0.001** |
| **log($f$)** $\times$ $\mathbf{1}\{f_{\geq 0.10}\}$ | 0.25 | 0.24 – 0.25 | **<0.001** |
| **log($n_{cases}$)** $\times$ $\mathbf{1}\{n_{cases \geq 30}\}$ | 0.14 | 0.14 – 0.15 | **<0.001** |
| Observations | 40874 | | |
| $R^2$ / $R^2$ adjusted | 0.727 / 0.727 | | |

**Table S3: Univariable model of coverage (equation 3 of section 1.5).**

| ***Predictors*** | ***Odds ratios*** | ***Confidence interval*** | ***p-value*** |
|---|---|---|---|
| (Intercept) | 32.20 | 32.00 – 32.39 | **<0.001** |
| $\boldsymbol{\sigma}_{\mathbf{peak}}$ | 0.78 | 0.78 – 0.78 | **<0.001** |
| Observations | 40874 | | |
| McFadden $R^2$ / McFadden $R^2$ adjusted | 0.182 / 0.182 | | |

**Table S4: Univariable model of specificity (equation 4 of section 1.5).**

| ***Predictors*** | ***Odds ratios*** | ***Confidence interval*** | ***p-value*** |
|---|---|---|---|
| (Intercept) | 36.73 | 36.41 – 37.06 | **<0.001** |
| $\boldsymbol{\sigma}_{\mathbf{peak}}$ | 0.76 | 0.76 – 0.76 | **<0.001** |
| Observations | 20396 | | |
| McFadden $R^2$ / McFadden $R^2$ adjusted | 0.236/0.236 | | |

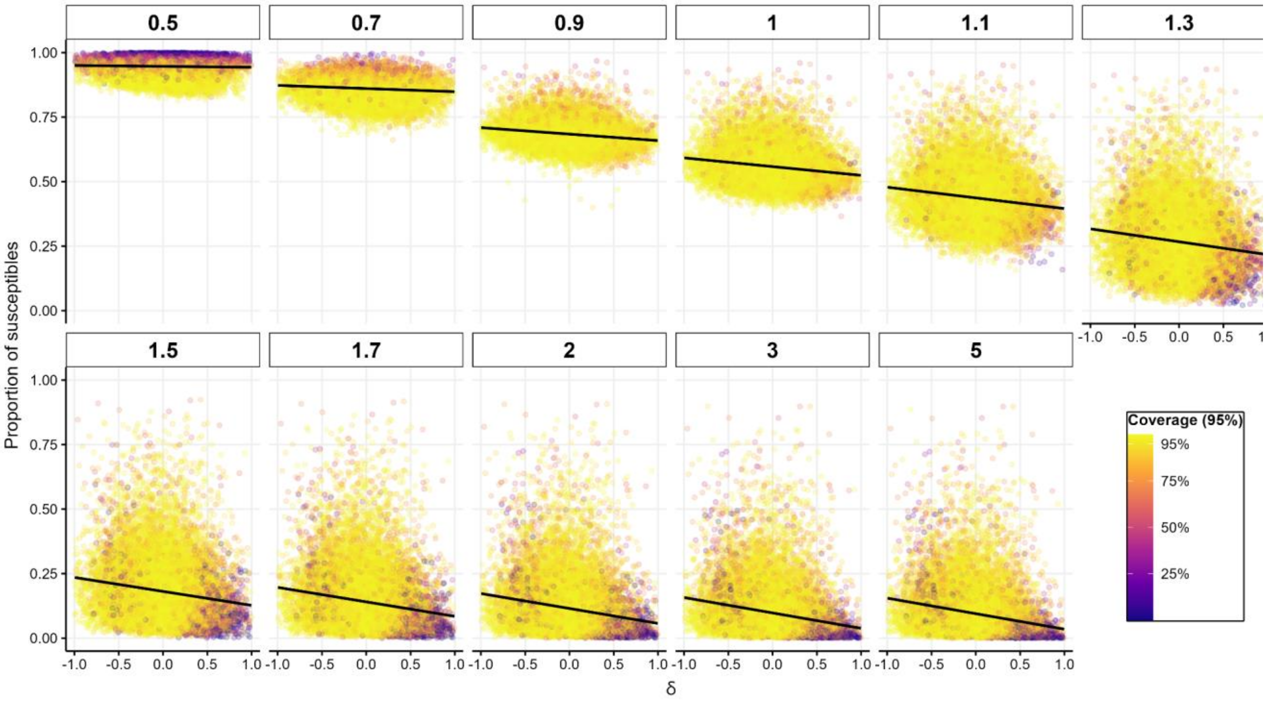


**Figure S4: Relationship between the assortativity coefficient of a group $\delta$ (x-axis), the proportion of susceptibles in that group (y-axis), and the 95% coverage of our estimator for $\delta$ (colour) across different epidemic stages (panels).**

Panel headers represent peak coefficient values. The black line depicts the linear regression between the group's proportion of susceptibles and its assortativity coefficient (excluding groups simulated with $\delta = 0$).

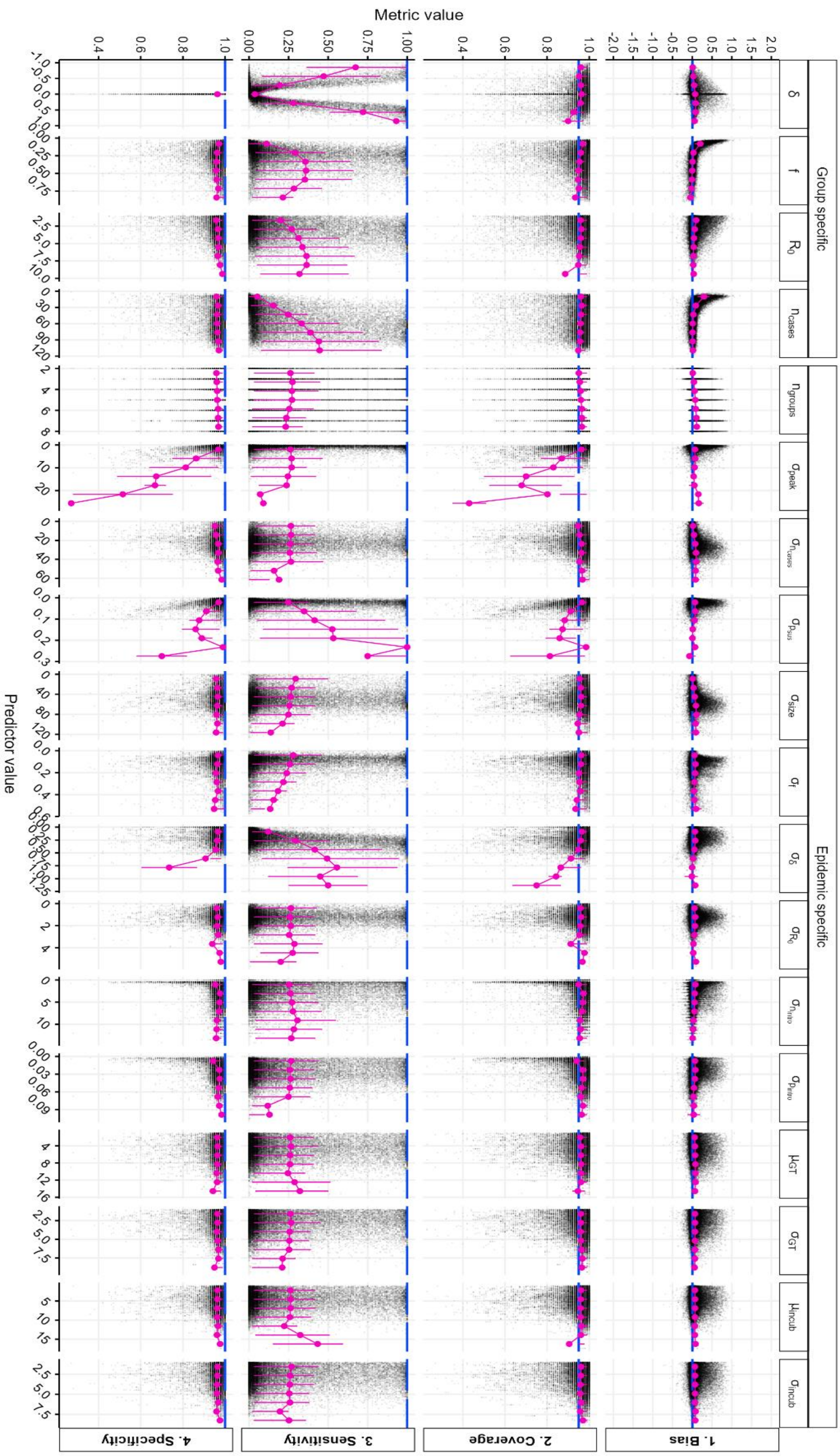
Metric value
Predictor value
Group specific
Epidemic specific
1. Bias
2. Coverage
3. Sensitivity
4. Specificity

**Figure S5: Estimator's performance across scenario parameters and epidemic characteristics.**
Figure 2 in the main text is a subset of this figure.
Each row corresponds to one performance indicator and each column corresponds to one simulation parameter or epidemic characteristic. In each panel, the scatter plot depicts the univariate relationship between simulation parameter or epidemic characteristic (x-axis) and the performance metric (y-axis), where each black dot represents the average observation from 100 simulations for each group in every scenario. The pink points and error bars indicate the mean and interquartile range, computed for all predictors (columns) within seven equally sized intervals. Dashed blue lines indicate target metric value. Transmission chains were analysed up to the group's epidemic peak with a significance level of 0.05.
Labels are defined as follows:

- $\delta$ : the true δ value for the group.
- $f$: proportion of the population belonging to the group.
- $R_0$: basic reproduction number for the group .
- $n_{cases}$: number of cases in the group.
- $n_{groups}$: total number of groups.
- $\sigma_{peak}$: observed standard deviation of the peak dates across all groups ('peak asynchronicity').
- $\sigma_{ncases}$: observed standard deviation of the number of cases across all groups.
- $\sigma_f$: observed standard deviation of the relative groups' sizes.
- $\sigma_\delta$: standard deviation of the groups' assortativity coefficients.
- $\sigma_{R0}$: standard deviation of the groups' basic reproduction number.
- $\sigma_{pintro}$: standard deviation of the groups' proportion of introductions.
- $\mu_{GT}$: mean of the generation time distribution.
- $\sigma_{GT}$: standard deviation the generation time distribution.
- $\mu_{incub}$: mean of the incubation period distribution.
- $\sigma_{incub}$: standard deviation in the incubation period distribution.

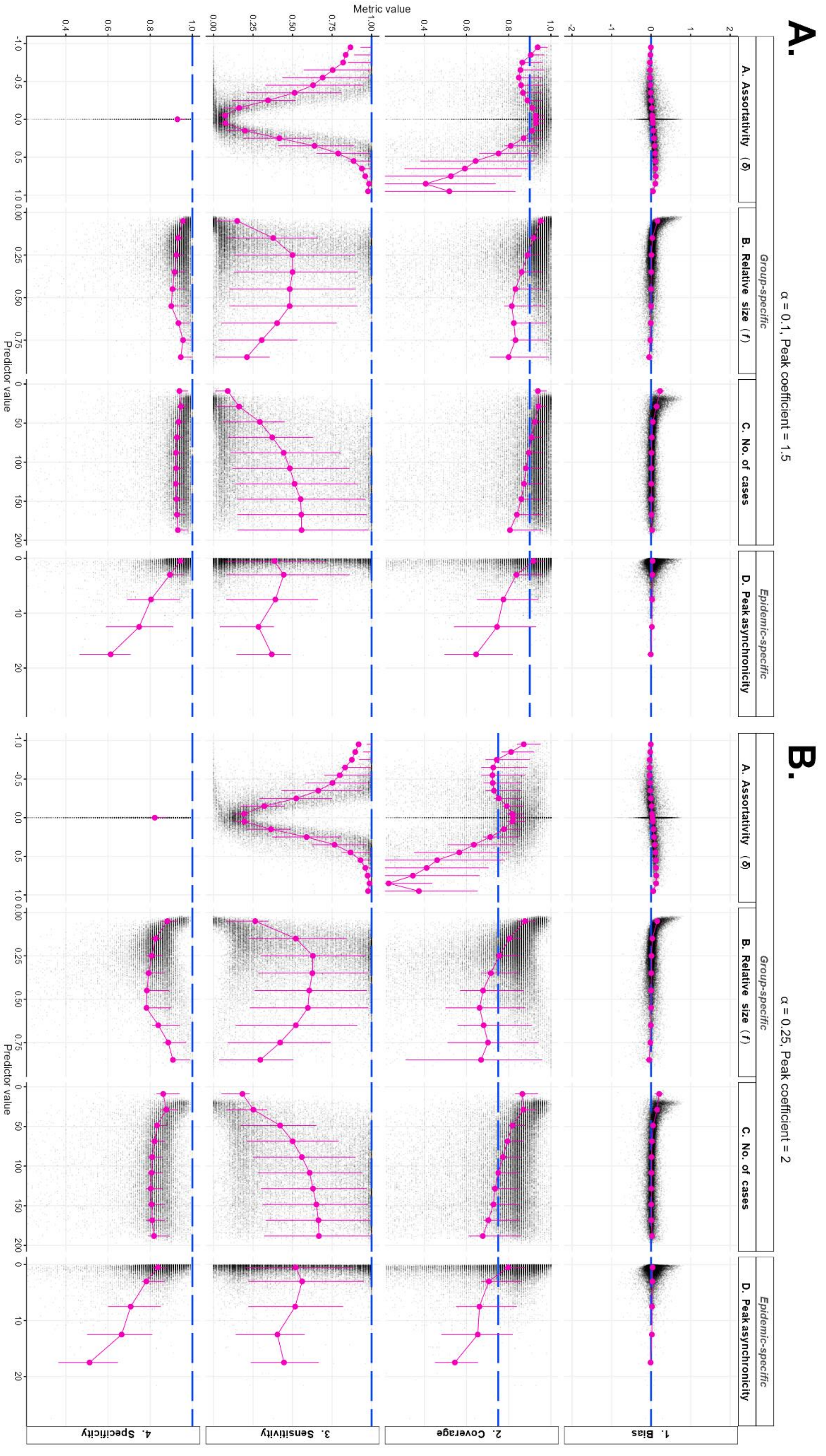
A.
α = 0.1, Peak coefficient = 1.5
B.
α = 0.25, Peak coefficient = 2
Group-specific
Epidemic-specific
A. Assortativity (δ)
B. Relative size (f)
C. No. of cases
D. Peak asynchronicity
1. Bias
2. Coverage
3. Sensitivity
4. Specificity
Metric value
Predictor value

**Figure S6: Estimator's performance across scenario parameters and epidemic characteristics. Same as main text Figure 2, but where:** A. Transmission chains have been analysed up to halfway after the group's epidemic peak (ε = 1.5) with a significance level of 0.1.

B. Transmission chains have been analysed using a peak coefficient ε = 2 with a significance level of 0.25.
